# Increasing incidence of parosmia and phantosmia in patients recovering from COVID-19 smell loss

**DOI:** 10.1101/2021.08.28.21262763

**Authors:** Kathrin Ohla, Maria Geraldine Veldhuizen, Tomer Green, Mackenzie E. Hannum, Alyssa J. Bakke, Shima T. Moein, Arnaud Tognetti, Elbrich M. Postma, Robert Pellegrino, Liang-Dar Hwang, Javier Albayay, Sachiko Koyama, Alissa Nolden, Thierry Thomas-Danguin, Carla Mucignat-Caretta, Nick S. Menger, Ilja Croijmans, Lina Öztürk, Hüseyin Yanık, Denis Pierron, Veronica Pereda-Loth, Alexia Nunez-Parra, Aldair M. Martinez Pineda, David Gillespie, Michael C. Farruggia, Cinzia Cecchetto, Marco A. Fornazieri, Carl Philpott, Vera Voznessenskaya, Keiland Cooper, Paloma Rohlfs Dominguez, Orietta Calcinoni, Jasper de Groot, Sanne Boesveldt, Surabhi Bhutani, Elisabeth M. Weir, Cara Exten, Paule V. Joseph, Valentina Parma, John E. Hayes, Masha Y. Niv

## Abstract

**Importance:** Sudden smell loss is a specific early symptom of COVID-19, with an estimated prevalence of ~40% to 75%. Smell impairment affects physical and mental health, and dietary behavior. Thus, it is critical to understand the rate and time course of smell recovery.

**Objective:** To characterize smell function and recovery up to 11 months post COVID-19 infection.

**Settings, Participants:** This longitudinal survey of individuals suffering COVID-19-related smell loss assessed disease symptoms and gustatory and olfactory function. Participants (n=12,313) who completed an initial respiratory symptoms, chemosensory function and COVID-19 diagnosis survey (S1) between April and September 2020 and completed a follow-up survey (S2) between September 2020 and February 2021; 27.5% participants responded (n=3,386), with 1,468 being diagnosed with COVID-19 and suffering co-occurring smell and taste loss at the beginning of their illness.

**Main Outcomes & Measures:** Primary outcomes are ratings of smell and taste function on a visual analog scale, and self-report of parosmia (smell distortions) and phantosmia (unexplained smells). Secondary outcomes include a checklist of other COVID-19 symptoms.

**Results:** On follow-up (median time since COVID-19 onset ~200 days), ~60% of women and ~48% of men reported less than 80% of their pre-illness smell ability. Taste typically recovered faster than smell, and taste loss rarely persisted if smell recovered. Prevalence of parosmia and phantosmia was ~10% of participants in S1 and increased substantially in S2: ~47% for parosmia and ~25% for phantosmia. Persistent smell impairment was associated with more symptoms overall, suggesting it may be a key marker of long-COVID. During COVID-19 illness, the ability to smell was slightly lower among those who did not recover their pre-illness ability to smell at S2.

**Conclusions and Relevance:** While smell loss improves for many individuals who lost it due to COVID-19, the prevalence of parosmia and phantosmia increases substantially over time. Olfactory dysfunction is also associated with wider COVID-19 symptoms and may persist for many months after COVID-19 onset. Taste loss in the absence of smell loss is rare. Persistent qualitative smell symptoms are emerging as common long term sequelae; more research into treatment options is strongly warranted given that conservative estimates suggest millions of individuals may experience parosmia following COVID-19. Healthcare providers worldwide need to be prepared to treat post COVID-19 secondary effects on physical and mental health.

**Trial registration:** This project was pre-registered at OSF: https://osf.io/3e6zc.

**Graphical abstract:** 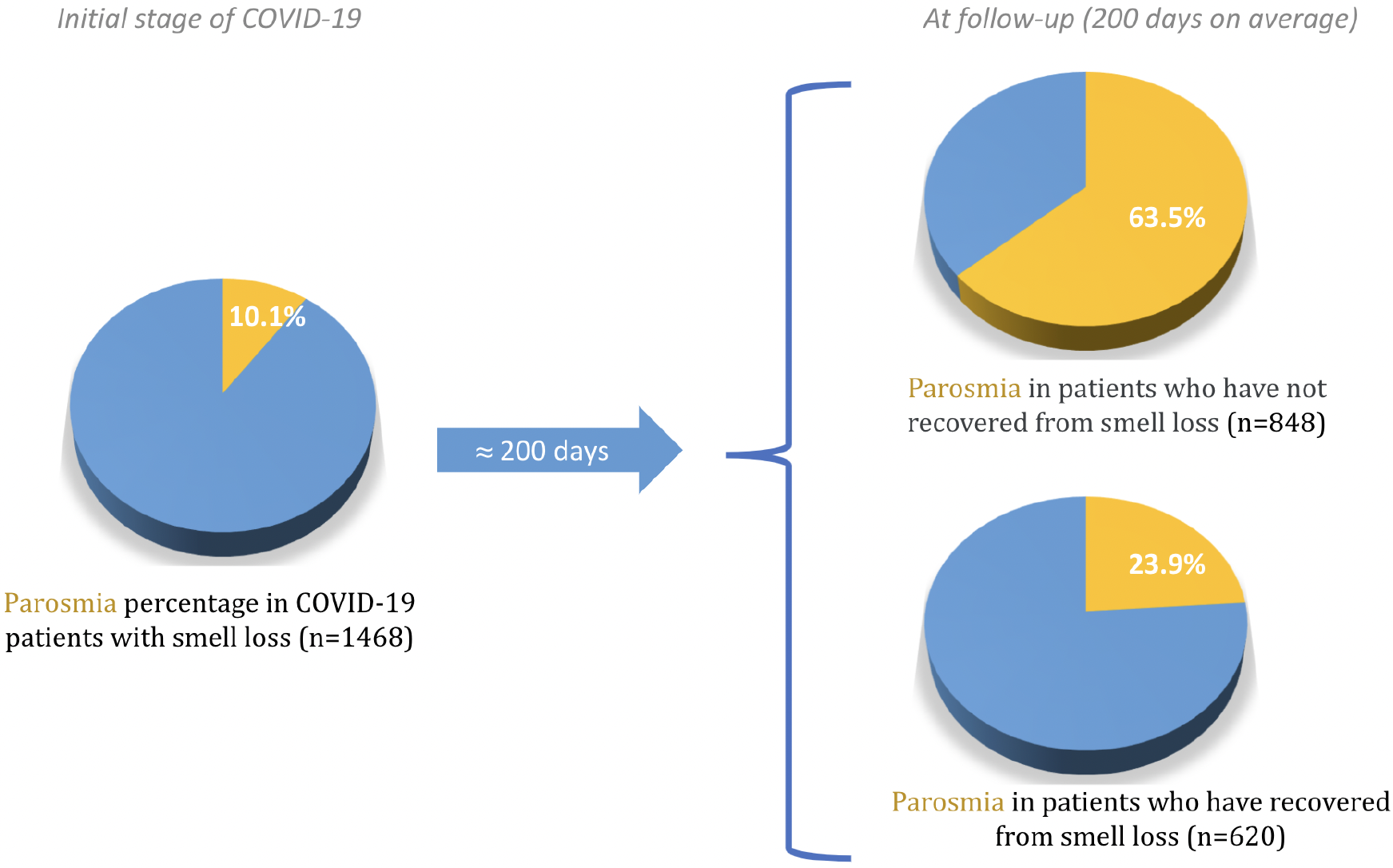

**Key Points:** *Question:* What are the characteristics of smell and taste recovery of COVID-19 patients?

*Findings:* In this preregistered observational study of 1,468 participants, smell loss is associated with a higher number of COVID-19 symptoms, and may persist for at least 11 months following disease onset. While a majority of participants report quantitative improvement in their ability to smell, the prevalence of parosmia and phantosmia increases substantially at follow-up. Taste recovers faster than smell, suggesting taste and smell recover separately and can be distinguished by the respondents.

*Meaning:* Olfactory dysfunction appears to be a component of long-COVID, with parosmia as a prominent symptom in almost half of those with smell loss. More research into treatment is needed, especially given that olfactory dysfunction is associated with depression and loss of appetite. Health professionals should be aware of these common and long lasting effects.

## 1. Introduction

### Background

In March 2020, the World Health Organization (WHO) declared that Coronavirus Disease 19 (COVID-19), caused by SARS-CoV-2 infection, had reached pandemic levels. Although the symptoms of COVID-19 are highly variable across infected individuals ^1^, sudden loss of taste and smell was quickly identified as a hallmark symptom ^2–4^. Self-reported smell loss was shown to be useful for both diagnosis ^5–7^ and population surveillance ^8^, at least for SARS-CoV-2 variants common in 2020.

Classically, patient complaints of smell loss with the common cold arise from a blocked or stuffy nose that prevents volatile odorants from reaching olfactory receptors near the top of the nasal cavity, and gustation is not affected ^9^. However, with COVID-19, sudden smell loss was commonly observed without nasal blockage ^10–12^, and direct assessment with odor-free tastants (e.g., sugar) indicated taste was also affected ^13^.

Most individuals (>75-80%) reporting taste and smell impairments due to COVID-19 tend to recover these senses within a few months, but smell impairment is still reported by 25-40% of patients after one or two months ^5,14^ at 6 months ^15,16^. Given the common confusion between taste, smell and flavor, data on taste recovery are less clear, though suggested to recover somewhat faster than smell ^16^. Separately, some individuals recover from acute smell loss, only to subsequently report other olfactory dysfunction, such as parosmia (smell distortions) and phantosmia (phantom smells or olfactory hallucinations) ^17,18^.

Factors associated with persistent smell and taste dysfunction remain unknown. Some early reports suggested smell loss might be associated with a milder disease course ^19,20^, although smell and taste impairments were also seen in severely ill patients ^21,22^.

### Objectives

The aim of this preregistered study (https://osf.io/3e6zc) was to characterize smell impairment and recovery in connection with taste loss and other symptoms, by recontacting respondents of our initial survey ^3,5^ to collect longitudinal data in a large cohort of participants diagnosed with COVID-19.

## 2. Methods

### Study design

This longitudinal, observational online cohort study entails a follow-up survey (S2) of respondents to the GCCR core survey (S1; https://www.nlm.nih.gov/dr2/COVID-19_BSSR_Research_Tools.pdf) ^3,5^ between 2 and 10 months after initial participation.

### Setting

Participants self-selected to participate in S1. They were invited via email to participate in S2 if they previously agreed to be re-contacted, provided an email address, completed S1 in English, Spanish, Italian, Dutch, French, and reported a change in smell, taste and/or flavor (via symptom checkbox) in S1. The protocol complies with the revised Declaration of Helsinki and was approved as an exempt study by the Office of Research Protections at The Pennsylvania Study University in the U.S.A. (STUDY00014904).

### Participants

#### Eligibility criteria

To be included in the present analysis, participants had to report a consistent COVID-19 diagnosis for both S1 and S2 – i.e., positive COVID-19 diagnosis via clinical presentation (i.e., via symptoms and history), or via viral swab, or another laboratory test. Other exclusion criteria are summarized in Figure 1.

**Figure 1.**
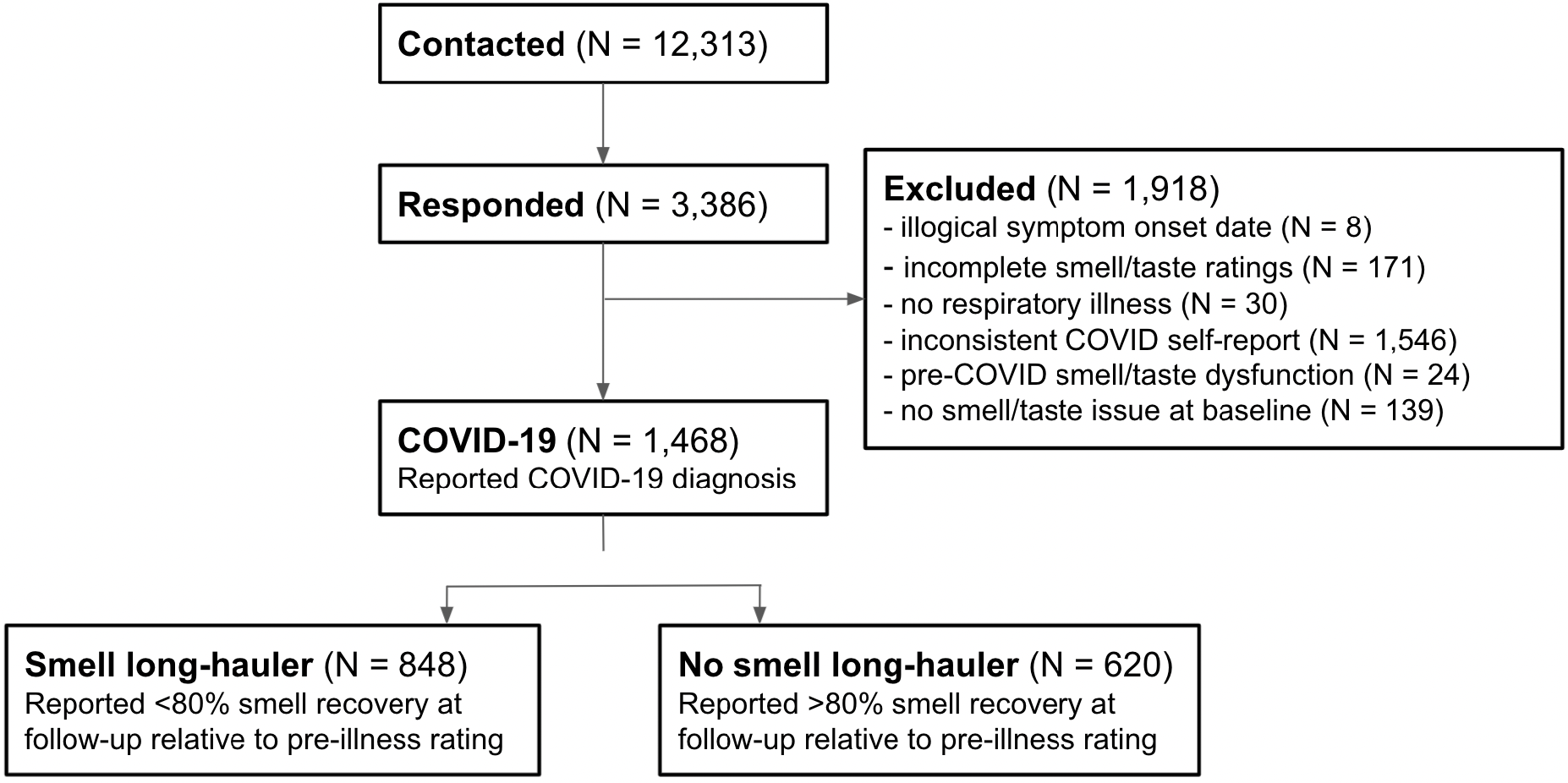
Summary of participants described in the current study. As shown in the exclusion box, the majority of S2 respondents were excluded from present analyses due to inconsistent reports of their COVID-19 diagnosis between S1 and S2. Participants were also excluded for missing or inconsistent data, chemosensory dysfunction prior to COVID-19.

### Variables, Data Sources and Measurement

Details of the baseline variables have been described previously ^3^. The follow-up survey collected ratings of smell and taste function on a visual analog scale, and self-reporting of parosmia and phantosmia. Other COVID-19 symptoms were collected via checklist and free text comments. Duplicate entries were removed.

### Bias minimization

The survey was conducted in multiple languages to increase generalizability. Also, because participants self-selected to respond to follow-up, analysis and conclusions were restricted to individuals with COVID-19 who had chemosensory loss at disease onset. As previously noted ^5^, focusing on this group allows for a conservative estimate of the relationship between smell dysfunction and COVID-19 diagnosis. Also, we excluded individuals with chemosensory impairment prior to COVID-19.

### Study size

There was no predetermination of the study size. A pilot inquiry in English only (n=100) was used to estimate feasible response rate among S1 completers ^23^, and invitations were sent out in the 5 languages with the greatest number of responses.

### Quantitative variables

Here, S2 respondents were grouped according to whether their smell loss persisted or recovered. Participants who returned to less than 80% of their pre-COVID smell ability (as reported in S1) were categorized as *smell long-haulers*; the rest were classified as *non long-haulers*. Smell (taste) impairment for the two surveys were calculated for each participant using the following equations

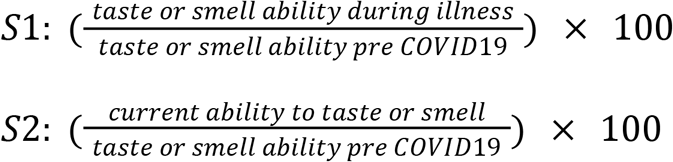

The percentage of individuals with smell and taste impairments was calculated relative to the total sample (N=1,468).

### Statistical Analysis

#### Demographics

To report demographics across the whole sample and to assess potential confounding variables, we calculated proportions of the presence of each of the following comorbidities: high blood pressure, heart disease, diabetes, obesity, lung disease (asthma/COPD), head trauma, neurological disease, cancer (treated with chemotherapy), cancer (no chemotherapy), chronic sinus problems, seasonal allergies/hay fever, and no condition. We also calculated the probability in each of the smell long-hauler groups. We tested distributional differences with Pearson’s chi-square tests with the R base function “prop.test”. We used an alpha of 0.0042 to determine significance (i.e., a Bonferroni corrected alpha of 0.05 for 12 conditions). We repeated this for language and gender distributions. For age we calculated the average and performed an independent sample t-test with an alpha of 0.05.

#### Differences in probability of smell distortions and other COVID-19 symptoms between participants with persistent versus recovered smell loss

To test differences in smell distortions *at the time of S2* between smell long-haulers and non long-haulers, we calculated probability tables of presence and absence of parosmia and phantosmia in each of the smell long-hauler groups. We tested distributional differences with Pearson’s chi-square tests with the R base function “prop.test”. We used an alpha of 0.025 to determine significance (i.e., a Bonferroni corrected alpha of 0.05 for two types of distortion). We repeated this analysis for the symptoms *at the time of S1* to check for any pre-existing differences prior to developing persistent smell long-hauler status.

#### Differences in symptom counts

To assess effects of smell long-hauler-status on illness severity, we summed the presence of each of commonly listed COVID-19 symptoms (fever, dry cough, cough with mucus, difficulty breathing / shortness of breath, chest tightness, runny nose, sore throat, loss of appetite, headache, muscle aches, fatigue, diarrhea, abdominal pain, nausea, excluding smell and taste symptoms under “changes in food flavor” and “changes in smell”), leading to scores ranging from 0-14. Since this “count” variable was not continuous or categorical (i.e., the total number of symptoms), we used logistic regression with a Poisson distribution for the dependent variable. This was implemented via the “glm” function in R, using the “poisson” option. The assumption of equality between variance and mean of each category of the independent variable was checked ^24^ and a “quasi-Poisson” family variant was applied if overdispersion was observed. To estimate relative risk, a Poisson regression with a robust error variance was calculated with the package Sandwich ^25–27^.

To further characterize rare symptoms not provided in the COVID-19 symptoms checklist, additional symptoms, such as “brain fog”, “memory loss”, were extracted from free text comments. Comments in Spanish, Italian, Dutch, and French were translated into English by scientists who were native speakers of each language, and pooled. In total, 559 comments containing symptoms were analysed [214 French (74 men, 140 women), 195 English (54 men, 141 women), 65 Spanish (22 men, 43 women), 54 Dutch (14 men, 40 women), and 31 Italian (13 men, 18 women)].

To test for differences in overall symptoms between smell long-haulers and non long-haulers at S2, we calculated probabilities for each of the 16 symptoms (headache, fatigue, difficulty breathing/shortness of breath, diarrhoea, nausea, fever, abdominal pain, changes in food flavour, changes in smell, chest tightness, cough with mucus, dry cough, loss of appetite, muscle aches, runny nose, sore throat) in each group. As above, we tested for distribution differences, with a Bonferroni corrected alpha of 0.003125 (0.05/16 tests, one for each symptom). We repeated this analysis for S1 symptoms to check for preexisting differences prior to developing smell long-hauler status.

Smell ability during COVID-19 infection (measured at S1) was compared between smell long-haulers and non long-haulers (defined from S2) using a Welch’s test.

## 3. Results

### Participants

Participants (n=12,313) who completed the initial online survey (S1) between April and September 2020 and agreed to be recontacted via email were invited to complete a follow-up survey (S2). Email invitations were sent in five languages (English: n=3,422, Spanish: n=1,575, Italian: n=1,165, Dutch: n=1,840, French: n=4,306) between September and November 2020 to those who consented to be re-contacted. Data were exported in February 2021. We received 3,386 responses (2,448 women, 927 men, 1 non-binary; age range 20 to 85 years) for S2, a response rate of ~28%. Of these, 1,918 participants were excluded from further analysis (see Figure 1 for details). Thus, the final dataset reported here consisted of 1,468 responses from individuals who reported smell or taste loss at baseline (S1) and reported consistent positive COVID-19 diagnoses at S1 and S2. The demographics, and overall symptoms of these individuals are reported in Table 1.

**Table 1.** Descriptive data of all participants and the smell long-hauler (LH) and no smell long-hauler (nLH) groups.

|  |  | All |  | LH |  | nLH |  | statistics |  |  |  |  |
| --- | --- | --- | --- | --- | --- | --- | --- | --- | --- | --- | --- | --- |
| Categorical variables |  | % | n | % | n | % | n | Chi2 | p | OR | CI lo | CI hi |
| **Prior conditions** |  |  |  |  |  |  |  |  |  |  |  |  |
|  | High blood pressure | 8.24 | 121 | 7.67 | 65 | 9.03 | 56 | 0.71 | 0.398 | 0.84 | -0.14 | 0.05 |
|  | Heart disease | 0.41 | 6 | 0.59 | 5 | 0.16 | 1 | 0.73 | 0.392 | 3.67 | -0.13 | 0.64 |
|  | Diabetes | 2.04 | 30 | 1.89 | 16 | 2.26 | 14 | 0.10 | 0.757 | 0.83 | -0.24 | 0.15 |
|  | Obesity | 8.99 | 132 | 9.79 | 83 | 7.90 | 49 | 1.33 | 0.248 | 1.26 | -0.03 | 0.15 |
|  | Lung disease (asthma / copd) | 5.11 | 75 | 5.31 | 45 | 4.84 | 30 | 0.08 | 0.778 | 1.10 | -0.10 | 0.14 |
|  | Head trauma | 0.14 | 2 | 0.24 | 2 | 0.00 | 0 | 0.24 | 0.621 | Inf | 0.15 | 0.70 |
|  | Neurological disease | 0.68 | 10 | 0.94 | 8 | 0.32 | 2 | 1.23 | 0.268 | 2.94 | -0.08 | 0.52 |
|  | Cancer (chemotherapy) | 0.14 | 2 | 0.12 | 1 | 0.16 | 1 | 0.00 | 1.000 | 0.73 | -0.85 | 0.69 |
|  | Cancer (no chemotherapy) | 0.14 | 2 | 0.24 | 2 | 0.00 | 0 | 0.24 | 0.621 | Inf | 0.15 | 0.70 |
|  | Chronic sinus problems | 4.02 | 59 | 3.42 | 29 | 4.84 | 30 | 1.52 | 0.218 | 0.70 | -0.23 | 0.05 |
|  | Seasonal allergies/hay fever | 16.42 | 241 | 16.27 | 138 | 16.61 | 103 | 0.01 | 0.919 | 0.98 | -0.08 | 0.06 |
|  | No conditions | 60.15 | 883 | 59.79 | 507 | 60.65 | 376 | 0.08 | 0.781 | 0.96 | -0.06 | 0.04 |
| **Gender** |  |  |  |  |  |  |  | 18.87 | 7.98E-05 |  |  |  |
|  | women | 75.68 | 1111 | 60.85 | 676 | 39.15 | 435 |  |  |  |  |  |
|  | men | 24.25 | 356 | 48.03 | 171 | 51.97 | 185 |  |  |  |  |  |
|  | non-binary | 0.07 | 1 | 100.00 | 1 | 0.00 | 0 |  |  |  |  |  |
| **Language** |  |  |  |  |  |  |  | 30,30 | 4,26E-06 |  |  |  |
|  | Dutch | 9.13 | 134 | 64.93 | 87 | 35.07 | 47 |  |  |  |  |  |
|  | English | 37.74 | 554 | 62.27 | 345 | 37.73 | 209 |  |  |  |  |  |
|  | French | 33.65 | 494 | 47.98 | 237 | 52.02 | 257 |  |  |  |  |  |
|  | Italian | 6.81 | 100 | 66.00 | 66 | 34.00 | 34 |  |  |  |  |  |
|  | Spanish | 12.67 | 186 | 60.75 | 113 | 39.25 | 73 |  |  |  |  |  |
| **Age in years** |  | **mean** | **SD** | **mean** | **SD** | **mean** | **SD** | **t-test** | **p** |  | **CI lo** | **CI hi** |
|  |  | 43.89 | 12.17 | 44.37 | 12.16 | 43.23 | 12.18 | -1.76 | 0.078 |  | -2.40 | 0.13 |

### Descriptive data

Descriptive data for all 1,468 participants are summarized in Table 1. The mean age was ~44 years, fewer men than women took part,, and more responses were collected in English and French, as expected from the relative distribution of email invitations sent.

### Outcome data

Time elapsed between S1 and S2 ranged from 23 to 291 days (median: 200 days) (see Supplementary Figure S1), corresponding to 36 to 326 days (median: 225 days) since disease onset; this timing enabled the calculation of cumulative rate of recovery (Table 2).

**Table 2:** Cumulative percentage of participants who recovered a pre-illness ability to smell or taste by months from the onset of disease

| **Time in months** | 2 | 3 | 4 | 5 | 6 | 7 | 8 | 9 | 10 | 11 |
| --- | --- | --- | --- | --- | --- | --- | --- | --- | --- | --- |
| **Smell** |  |  |  |  |  |  |  |  |  |  |
| Women | 0.18 | 1.26 | 2.43 | 4.05 | 6.66 | 12.69 | 27.27 | 29.43 | 35.37 | 39.15 |
| Men | 0.28 | 1.68 | 3.93 | 6.17 | 9.26 | 14.88 | 35.95 | 37.92 | 45.22 | 51.96 |
| **Taste** |  |  |  |  |  |  |  |  |  |  |
| Women | 0.27 | 1.89 | 3.87 | 5.94 | 10.08 | 18.72 | 39.33 | 43.20 | 51.93 | 56.07 |
| Men | 0.56 | 2.24 | 4.77 | 7.30 | 11.23 | 18.53 | 45.78 | 48.31 | 58.70 | 64.88 |

### Main results

During the first months after onset of COVID-19 symptoms, less than 10% of participants reported full smell recovery, gradually increasing to 39% in women and 52% in men by up to 11 months (Table 2). Comparatively, the reports for taste recovery were greater (~56 to ~65% by 11 months).

58% of those in the final S2 dataset were classified as smell long-haulers (see methods), with ~39% also reporting persistent taste impairment and ~20% reporting recovered taste (Figure 2A). Only ~3% reported impaired taste with recovered smell. This suggests smell and taste recover separately, and these sensory abilities can be distinguished by the respondents.

**Figure 2.**
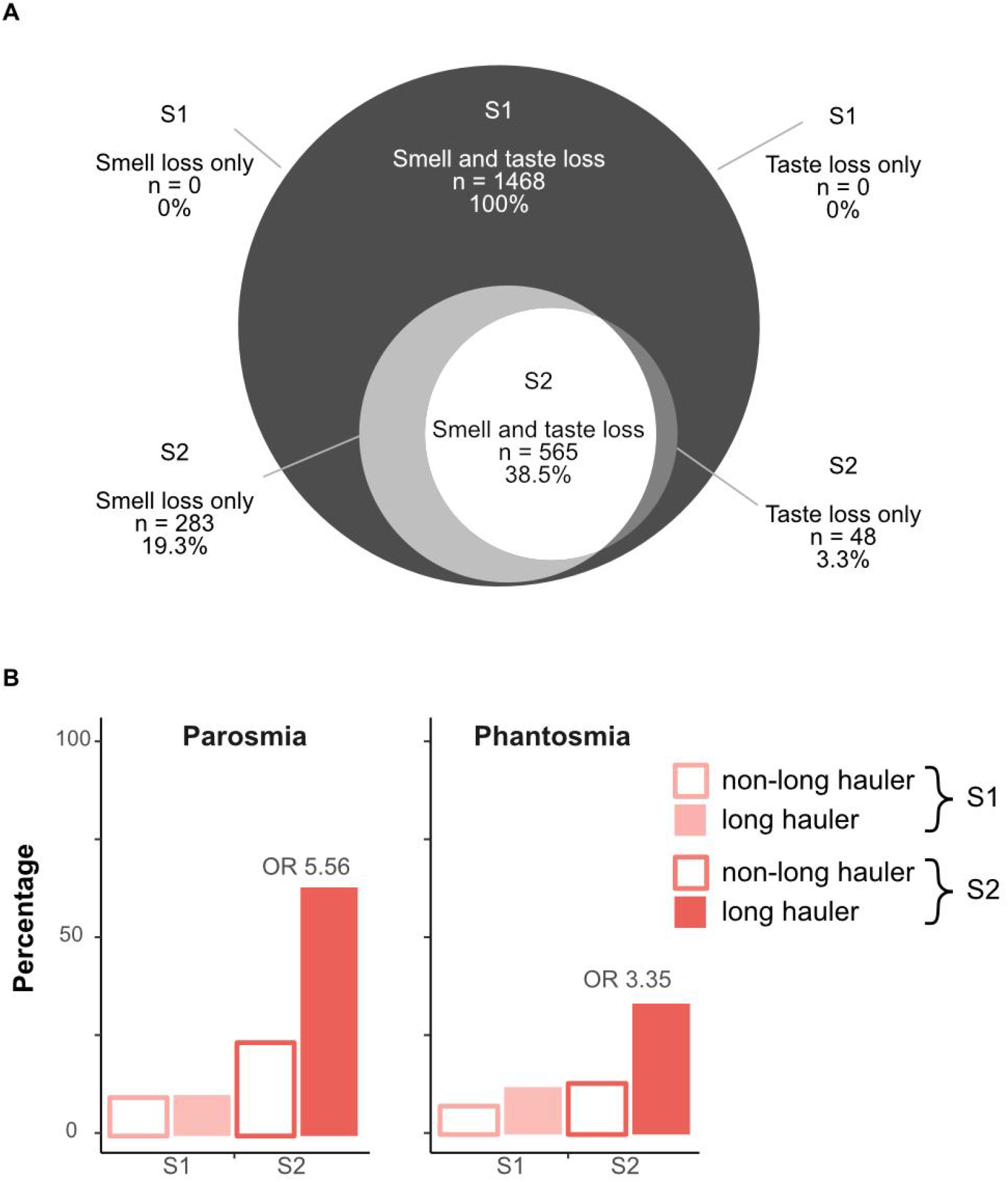
A: Proportions of participants with smell, taste, and combined smell and taste impairments during baseline (S1, dark gray) and follow-up (S2, lighter grays and white). B: S1 (light pink) and S2 (dark pink) proportions of qualitative smell changes – specifically, parosmia and phantosmia – for individuals who will regain smell ability (white fill) or exhibit smell long-hauling (solid fill).

Qualitative disorders of smell, specifically parosmia and phantosmia, were more frequently observed at S2 (46.8% and 24.7%, respectively) than S1 (10.2% and 10.1%, respectively; Figure 2B). Further, these types of dysfunction were significantly more common in smell long-haulers than in non-long-haulers, 63.6% of smell long-haulers reported parosmia versus 23.9% of non-long-haulers (χ^2^ = 225.0, 95% CI = 0.34-0.44, p <.001, OR = 5.56) and 33.5% of smell long-haulers reported phantosmia versus 13.1% of non-long-haulers (χ^2^ = 78.9, 95% CI = 0.21-0.32, p <.001, OR = 3.35). Among smell long-haulers, the incidence of parosmia was not significantly different between women and men (64% versus 58%). Qualitative terms from open-ended text responses were also captured. Typical participant reports for parosmia were “Some things now smell different and unpleasant” or “like chemicals”; reports for phantosmia include responses like “Sometimes I can smell burning but no one else around me can.”

The total number of symptoms decreased at S2 (see Figure 3). However, smell long-haulers reported more overall symptoms (median = 1) at S2 compared to non-long-haulers (median = 0). This was confirmed via quasi-Poisson regression (β_1_ = 0.48, 95% CI = 0.32-0.64, T = 5.66, p < .0001). Notably, these groups were not different at S1 (both medians = 6).

**Figure 3.**
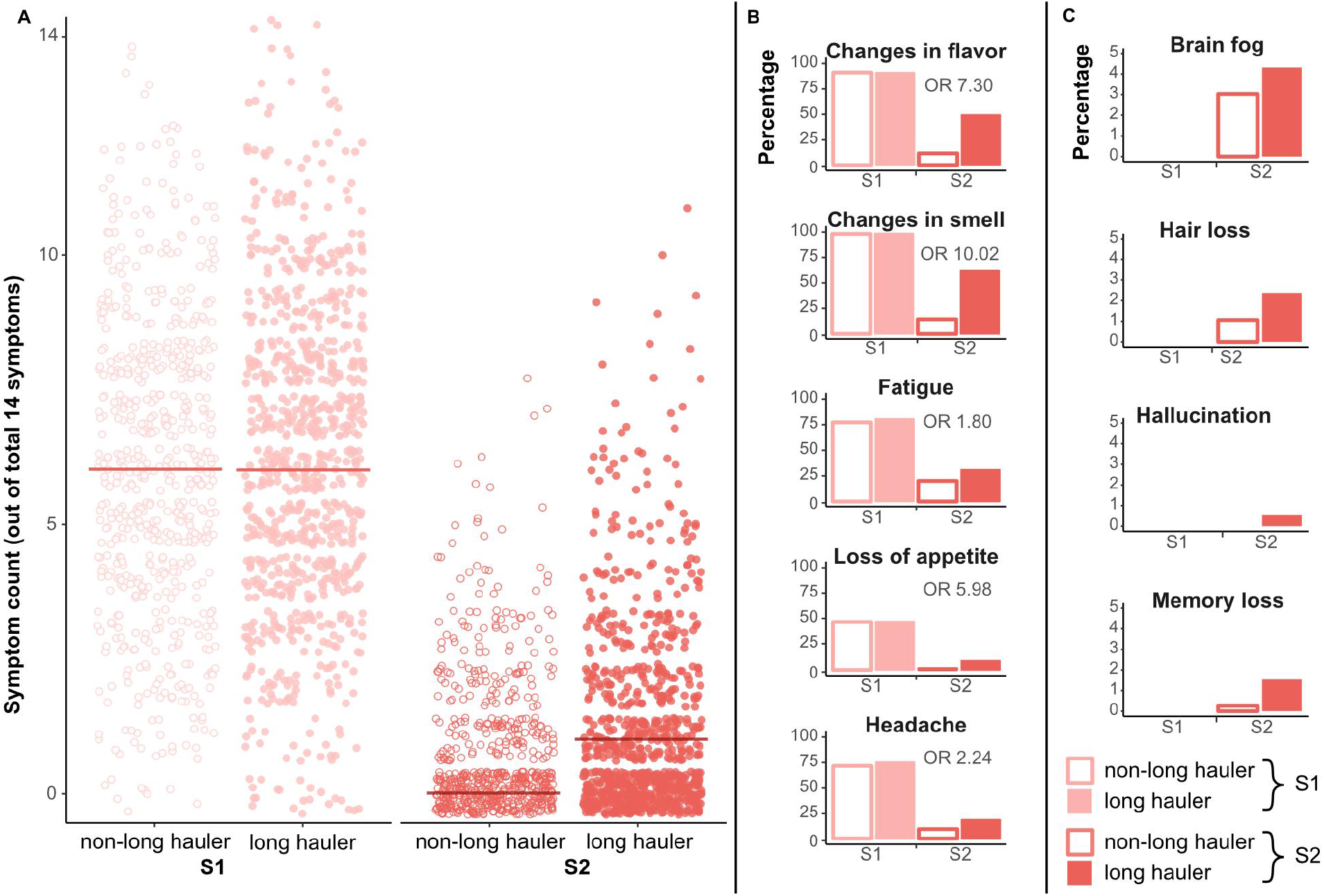
A: Comparison of overall number of non-chemosensory symptoms at baseline (S1, light pink) and follow-up (S2, dark pink), stratified by smell long-hauler (LH) status at S2. B: Comparison of selected symptoms at S1 and S2 stratified by smell long-haulers status at S2. C: Percentage of rare symptoms spontaneously mentioned in free text responses in English, Spanish, Dutch, Italian, and French.

When we examine each of the symptoms, including smell and taste symptoms, we observed changes in flavor (χ^2^ = 224.9, 95% CI = 0.37-0.46, p <.001, OR = 7.30) and in smell (χ^2^ = 340.17, 95% CI = 0.44-0.53, p <.001, OR = 10.02) as expected, in addition to other symptoms like fatigue (χ^2^ = 22.09, 95% CI = 0.08-0.20, p <.001, OR = 1.80), headache (χ^2^ = 23.99, 95% CI = 0.11-0.25, p <.001, OR = 2.24), and loss of appetite (χ^2^ = 33.58, 95% CI = 0.25-0.40, p <.001, OR = 5.98), all of which were more frequent in smell long-haulers than in non-long-haulers (Figure 3B). This suggests smell long-haulers had greater overall morbidity. Analysis of spontaneous mentions of rare symptoms in free text responses also supports the notion that smell long-haulers experience more symptoms: spontaneous comments included brain fog, hair loss, hallucination, and memory loss. Formal statistics were not applied due to low incidence of these reports (Figure 3C).

To identify variables with potential prognostic value in predicting who would eventually become a smell long-hauler, we looked for differences in multiple S1 measures across the smell long-hauler and non-long-hauler groups from S2. None of these were significant, save one: the self-rated ability to smell during COVID-19 illness was slightly lower (Welch’s t-test, statistic = −4.33, p <.0001) in smell long-haulers (n=848) than in non-long-haulers (n=620), with means of 2.96 (± 7.64, 95% CI = 2.45-3.48) and 5.11 (±10.49, 95% CI = 4.28-5.94), respectively. This was confirmed when the distributions of smell ability at S1 were compared by status at S2 (Kolmogorov–Smirnov test statistic = 0.12; p<0.0001). As shown in Figure 4, a greater number of smell long-haulers rated their smell ability during illness below 5 (on a 101-point scale), relative to non-long-haulers, although the prognostic value of this small difference still needs to be confirmed.

**Figure 4.**
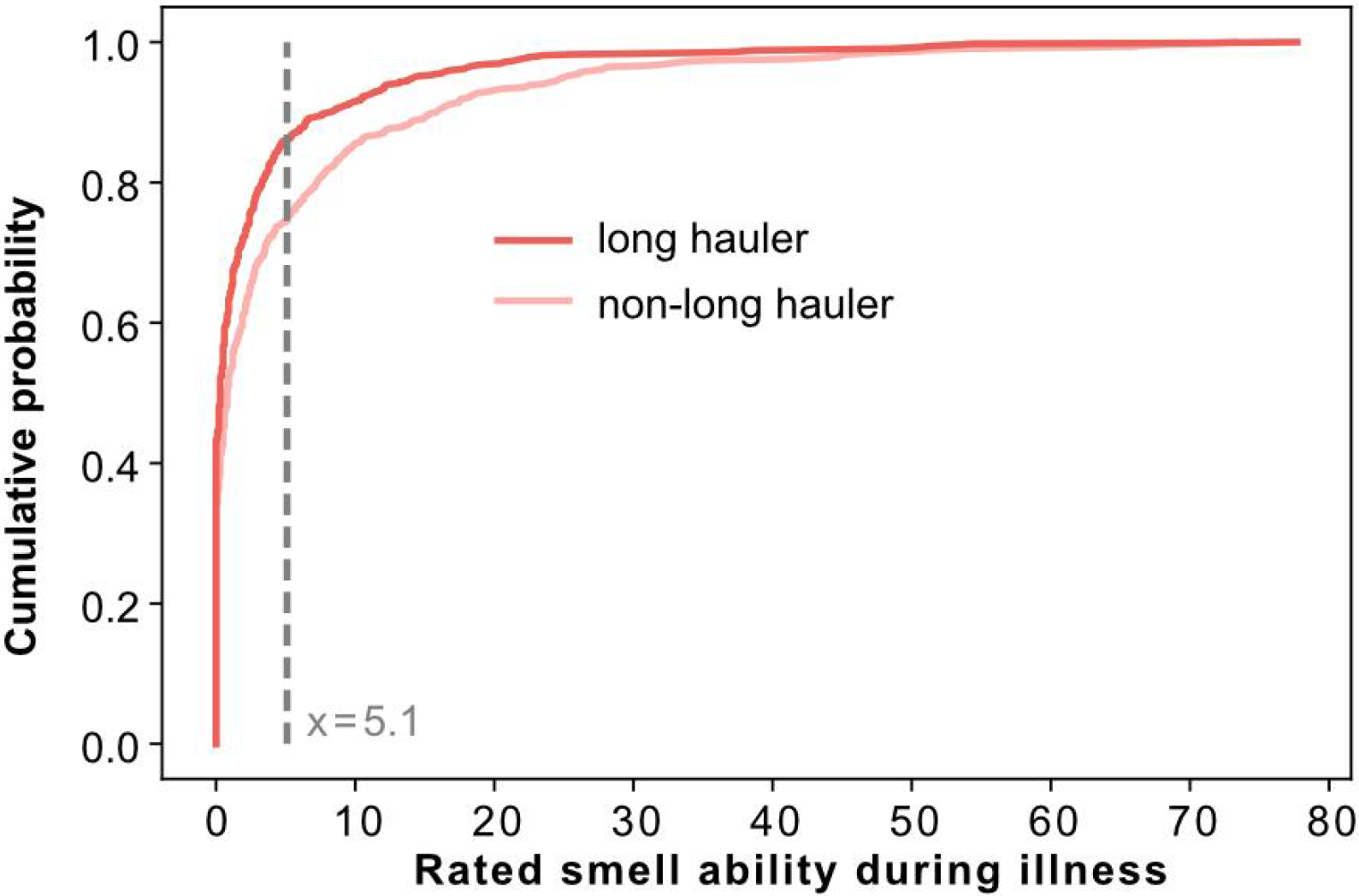
Distribution of ratings for smell ability at baseline (S1), stratified by whether a participant was classified as a smell long-hauler (dark pink) or non long-hauler (light pink) at follow-up (S2). At baseline, the majority of both groups (i.e., more than 50%) show complete smell loss (a score of zero on the x-axis), but a higher proportion of those who would later recover (light pink line) were hyposmic, rather than totally anosmic, at baseline. The dashed vertical line indicates the smell ability rating (on a VAS from 0-100) where the two groups differ maximally.

Given recent work on predictors of long-COVID ^28^, we also performed a supplementary analysis (see Supplement) on a sub-group of participants to compare the fully recovered (N=153) with those still experiencing at least one long-term symptom (N=202). The number of overall symptoms experienced during the first two weeks of the disease was predictive of having long-term symptoms. Consistent with ^28^, the greater the number of symptoms experienced by the participants during the first 2 weeks of the disease, the more likely they were to have long-term symptoms more than 2 months later. This is also in line with more severe outcomes of hospitalized vs non-hospitalized COVID-19 patients ^29^.

## 4. Discussion

### Key results

Our follow-up of 1482 participants suggests that ~60% of women and ~48% of men recover less than 80% of their pre-illness olfactory ability several months (200 days median) since COVID-19 onset. Taste recovered more quickly and rarely persisted if smell recovered. Prevalence of parosmia and phantosmia rose from 10% during the baseline survey to ~47% and ~25% at the follow-up. These olfactory dysfunctions were more common for smell long-haulers than non long-haulers. Persistent smell loss also coincided with more COVID-19 symptoms at follow up and a higher incidence of follow up symptoms, such as headache.

### Limitations

Participants for S1 were recruited via social media (with additional coverage in traditional media) for this web-based survey which may explain why participants under 60 years of age and women are overrepresented in our sample. The ~28% response rate for S2 may reflect that many S1 participants had spontaneously recovered olfactory and/or gustatory function and were therefore no longer interested in responding. The time lapse between disease onset and follow-up survey varies between participants. Furthermore, launch dates and pandemic situations varied between different countries, and time between surveys S1 and S2 differed by individuals. Despite these limitations, our findings characterize profiles of smell and taste loss recovery, with important downstream implications for public health.

### Interpretation

Previously, some speculated smell loss might indicate milder COVID-19 morbidity ^20^. Our data do not support this view; rather, we found smell long-haulers had more symptoms than recovered participants. This suggests previous under-reporting of smell dysfunction among severely ill patients may instead reflect a sampling bias; understandably it seems likely that clinicians treating critically ill patients were less focused on anosmia or parosmia as symptoms, and such patients were presumably unavailable for acute chemosensory testing.

There is important practical value in being able to predict which patients may develop long term smell loss. We found a greater reduction in the smell ability during COVID-19 illness in those who later became smell long-haulers compared to those who recovered smell ability, although this difference was numerically small. Whether the rating of smell ability during the disease can be used prognostically to predict the risk of future long-hauling needs additional exploration.

While some studies suggest self-reports may underestimate smell loss prevalence relative to direct assessment ^18,30,31^, others found correlations between self-reporting and direct assessments ^32^. Furthermore, although direct assessments have been proposed very recently ^33^, self-report remains the current standard of care for assessment of parosmia and phantosmia ^34^, at least until newly proposed methods can be further validated. The presence of parosmia in nearly 50% of the smell long-haulers in our sample is not surprising for post-viral olfactory dysfunction in general ^35^. In other datasets (i.e., UK healthcare workers), parosmia is also emerging as a common sequelae of COVID-19 ^36^.

### Generalizability

As of August 2021, there are over 36 million Americans and 210 million people worldwide recovering from COVID-19 ^37^. According to meta-analysis ^30^, 77% of those with COVID-19 have acute smell loss when smell function is measured directly or 44% if based on self-reports. If we conservatively assume half of those with COVID-19 experience acute smell loss, this suggests ~18 million Americans may have experienced acute anosmia. If we are highly conservative and assume all of the individuals who did not respond to our follow up survey recovered, we calculate 50% (smell long haulers) of 30% (response rate), resulting in ~2.7 million Americans and ~15 millions worldwide may be smell long-haulers. Present data suggest ~47% of smell long-haulers report parosmia, which would translate to over a million Americans (and over 7 million worldwide) with parosmia as a result of COVID-19.While olfactory symptoms may be formally classified as mild outcomes by some health authorities, the possibility that millions of individuals may experience long term anosmia and parosmia as a consequence of prior COVID-19 infection is highly concerning, given the downstream impacts this will likely have on dietary habits ^38^, quality of life ^39^, and mental health ^40^. We also find that smell long-haulers report other post acute sequelae of COVID-19.

## 5. Conclusion

Our study provides insights on the symptoms of a large number of individuals diagnosed with COVID-19, who experienced persistent smell and taste loss, up to 11 months (6-7 months median) since disease onset. Our finding that parosmia increases from ~10% at baseline to almost 50% at follow-up suggests parosmia may be a common symptom post-COVID-19. It is important that health providers, patients, and their families are aware of this potential development, and they are educated about the course of disease and management. Millions of people worldwide are likely affected and additional research as well as development of new treatment options are needed.

## Data Availability

Data will be made available upon acceptance of publication here: https://osf.io/e68ns/wiki/10_Projects/.

## Acknowledgments

We thank all members of the Global Consortium for Chemosensory Research (GCCR) for the core survey and for help with translations. We thank Danielle R. Reed and Thomas Hummel for their valuable contributions in the initial stages of this work. Deployment of the original GCCR survey and follow-up survey were both supported by an unrestricted gift from James and Helen Zallie to support sensory science research at Penn State.

## Supplement

**Supplementary Figure 1.**
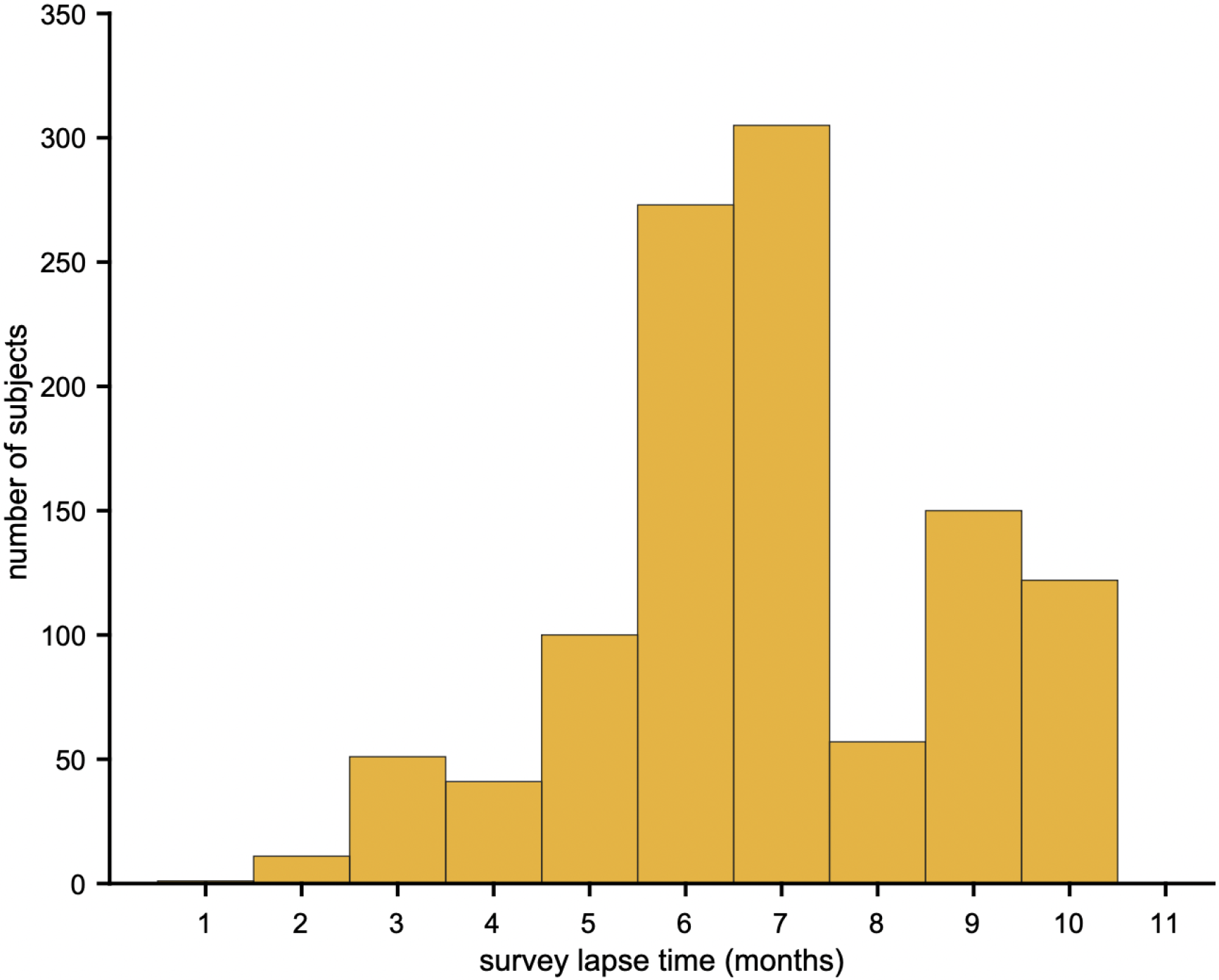
Distribution of time lapse between S1 and S2.

**Supplementary Table 1.** Comparison of symptoms between long haulers and non-longhaulers

| Survey | Symptom | longhauler |  | non long hauler |  | Chi-square | P-value | P-value Bonferroni corrected | Odds ratio | 95% CI |  |
| --- | --- | --- | --- | --- | --- | --- | --- | --- | --- | --- | --- |
|  |  | % | n | % | n |  |  |  |  | lower bound | higher bound |
| S1 | fever | 56.37% | 478 | 58.23% | 361 | 0.432 | 0.511 |  | 0.93 | -0.07 | 0.03 |
| S1 | dry cough | 54.48% | 462 | 56.45% | 350 | 0.486 | 0.486 |  | 0.92 | -0.07 | 0.03 |
| S1 | cough with mucus | 17.92% | 152 | 18.87% | 117 | 0.156 | 0.693 |  | 0.94 | -0.08 | 0.05 |
| S1 | difficulty breathing / shortness of brea | 37.85% | 321 | 36.94% | 229 | 0.093 | 0.761 |  | 1.04 | -0.04 | 0.06 |
| S1 | chest tightness | 36.79% | 312 | 32.42% | 201 | 2.824 | 0.093 |  | 1.21 | -0.01 | 0.10 |
| S1 | runny nose | 37.62% | 319 | 38.39% | 238 | 0.060 | 0.806 |  | 0.97 | -0.06 | 0.05 |
| S1 | sore throat | 38.09% | 323 | 33.71% | 209 | 2.787 | 0.095 |  | 1.21 | -0.01 | 0.10 |
| S1 | changes in food flavor | 91.04% | 772 | 91.13% | 565 | 0.000 | 1.000 |  | 0.99 | -0.09 | 0.09 |
| S1 | changes in smell | 98.70% | 837 | 98.23% | 609 | 0.276 | 0.599 |  | 1.37 | -0.15 | 0.31 |
| S1 | loss of appetite | 47.29% | 401 | 47.10% | 292 | 0.000 | 0.984 |  | 1.01 | -0.05 | 0.05 |
| S1 | headache | 75.00% | 636 | 71.29% | 442 | 2.340 | 0.126 |  | 1.21 | -0.01 | 0.11 |
| S1 | muscle aches | 63.56% | 539 | 60.81% | 377 | 1.044 | 0.307 |  | 1.12 | -0.03 | 0.08 |
| S1 | fatigue | 81.37% | 690 | 77.74% | 482 | 2.704 | 0.100 |  | 1.25 | -0.01 | 0.12 |
| S1 | diarrhea | 35.02% | 297 | 36.61% | 227 | 0.328 | 0.567 |  | 0.93 | -0.07 | 0.04 |
| S1 | abdominal pain | 18.40% | 156 | 14.68% | 91 | 3.278 | 0.070 |  | 1.31 | 0.00 | 0.13 |
| S1 | nausea | 26.42% | 224 | 22.26% | 138 | 3.111 | 0.078 |  | 1.25 | -0.01 | 0.11 |
| S2 | fever | 0.71% | 6 | 0.49% | 3 | 0.039 | 0.843 |  | 1.46 | -0.28 | 0.45 |
| S2 | dry cough | 6.49% | 55 | 4.38% | 27 | 2.640 | 0.104 |  | 1.52 | -0.01 | 0.21 |
| S2 | cough with mucus | 4.72% | 40 | 3.08% | 19 | 2.085 | 0.149 |  | 1.56 | -0.03 | 0.23 |
| S2 | difficulty breathing / shortness of brea | 10.39% | 88 | 6.81% | 42 | 5.228 | 0.022 | 0.356 | 1.59 | 0.02 | 0.20 |
| S2 | chest tightness | 6.26% | 53 | 6.32% | 39 | 0.000 | 1.000 |  | 0.99 | -0.11 | 0.10 |
| S2 | runny nose | 7.91% | 67 | 4.54% | 28 | 6.145 | 0.013 | 0.211 | 1.81 | 0.03 | 0.24 |
| S2 | sore throat | 4.25% | 36 | 3.08% | 19 | 1.049 | 0.306 |  | 1.40 | -0.06 | 0.22 |
| S2 | changes in food flavor | 49.47% | 419 | 11.83% | 73 | 224.940 | 0.000 | 0.000 | 7.30 | 0.37 | 0.46 |
| S2 | changes in smell | 62.81% | 532 | 14.42% | 89 | 340.170 | 0.000 | 0.000 | 10.02 | 0.44 | 0.53 |
| S2 | loss of appetite | 8.97% | 76 | 1.62% | 10 | 33.580 | 0.000 | 0.000 | 5.98 | 0.25 | 0.40 |
| S2 | headache | 18.54% | 157 | 9.24% | 57 | 23.990 | 0.000 | 0.000 | 2.24 | 0.11 | 0.25 |
| S2 | muscle aches | 11.69% | 99 | 8.27% | 51 | 4.180 | 0.041 | 0.654 | 1.47 | 0.01 | 0.17 |
| S2 | fatigue | 31.40% | 266 | 20.26% | 125 | 22.090 | 0.000 | 0.000 | 1.80 | 0.08 | 0.20 |
| S2 | diarrhea | 4.84% | 41 | 2.76% | 17 | 3.550 | 0.060 |  | 1.80 | 0.00 | 0.26 |
| S2 | abdominal pain | 4.96% | 42 | 4.38% | 27 | 0.156 | 0.693 |  | 1.14 | -0.09 | 0.16 |
| S2 | nausea | 5.43% | 46 | 3.24% | 20 | 3.483 | 0.062 |  | 1.71 | 0.00 | 0.25 |

### Differences in smell changes between COVID-19 antibody test results

Participants were asked whether they have been tested for the COVID-19 antibody at the follow-up. Among participants who had consistent COVID-19 diagnosis (1,471 positive and 913 negative), 1,064 and 203 reported having positive (Ab+) and negative (Ab-) antibody test results, respectively, with the remaining reporting no antibody test (n=1100) or unknown (n=17). We conducted a t-test to assess the difference in the self-report ability to smell between Ab+ and Ab- at four time points, which were before illness, during illness, most impaired and current. We showed that participants with Ab+ had lower ratings of smell during illness and at the most impaired period (Supplementary Figure 2). There were no differences before illness and at the current time. These results were consistent with our previous study comparing the ratings of smell between participants who were diagnosed with COVID-19 + and COVID-19 -, providing additional support for the reduced olfactory function in COVID-19.

**Supplementary Figure 2.**
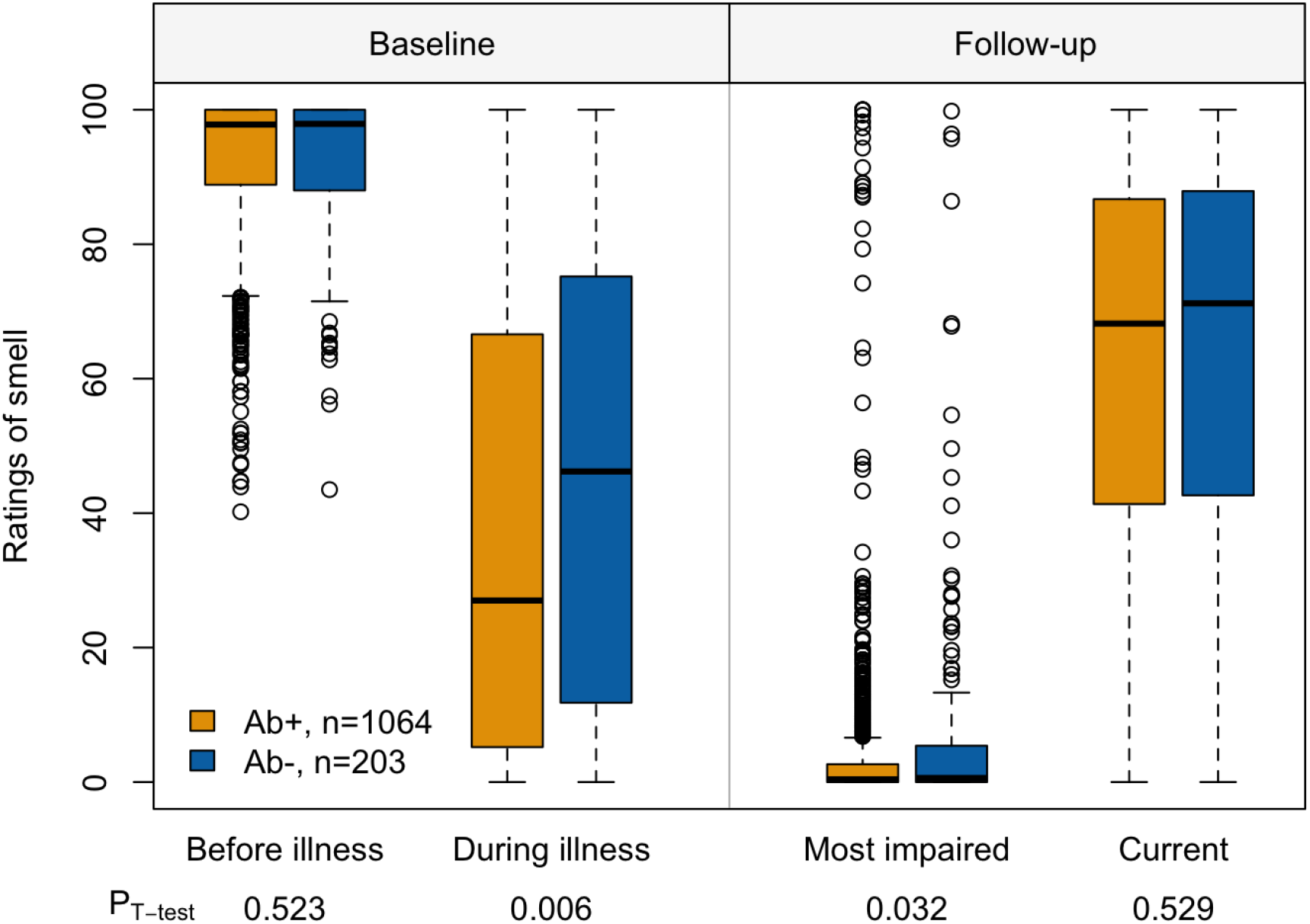
Comparison of self-report ability to smell between individuals who tested positive (Ab+) and negative (Ab-) for the COVID-19 antibody. Ratings before illness and during illness were collected at the baseline and most impaired and current ratings were collected at the follow-up.

### Is the number of symptoms experienced during the first two weeks of illness predictive of long-COVID?

#### Participants

To examine whether the number of symptoms experienced during the first two weeks of illness is predictive of long-COVID, we performed analyses on a separate sub-group of recontacted participants, namely COVID-19 positive participants who responded both to S1 during the first 14 days of illness and to S2 more than 2 months (≥61days) after disease onset. From this sub-group (N=355), we categorized participants according to their disease status at S2: those who reported to still experiencing at least 1 symptom more than 61 days after the disease onset were defined as ‘Long-COVID’ (N=202, 161 women, 41 men) while those who reported 0 symptom were defined as ‘Recovered’ (N=153, 104 women, 49 men).

#### Statistical analyses

We used a logistic regression (glm function with a binomial error structure of the stats package in R) to assess whether the two categories of participants (Long-COVID vs Recovered) differed in terms of overall number of symptoms they respectively experienced during the first two weeks of disease. Our dependent variable was the “Participants’ category” (Long-COVID vs Recovered). Our explanatory variable was the “Number of symptoms’’ reported during the first two weeks. We also included “Age” and “Gender” as control variables. Finally, we added the variable ‘Time-lapse’ corresponding to the number of days between disease onset and the date of S2 completion as a control variable. In other words, the model was: Participants’ category ~ Number of symptoms + Age + Gender + Time-lapse. We centred Age and Time-lapse in order to make the effects more easily biologically interpretable. The significance of each variable was tested with likelihood ratio tests comparing the full model to those without the term of interest and the α-level was set to 0.05.

## Results

The logistic regression revealed a significant effect of the number of symptoms during the first 14 days of disease (ß=0.10, SE = 0.04, 95% CI = 1.032-1.196, χ2 = 7.98, p =.005, OR = 1.11): participants who developed long-COVID (i.e., they are still experiencing at least one symptom after 61 days) experienced a significantly higher number of symptoms (Mean ± SD = 8.3 ± 3.07 symptoms) during the first 14 days of disease compared to the participants who had fully recovered after two months (Mean ± SD = 7.3 ± 2.87 symptoms). Importantly, the number of days between disease onset and S2 completion does not significantly differ between the two categories of participants (ß=-0.001, SE = 0.002, 95% CI = 0.995-1.003, χ2 = 0.24, p =.62, OR = 1.00). No significant effect of age (ß=0.02, SE = 0.009, 95% CI = 0.997-1.036, χ2 = 2.84, p =.09, OR = 1.02) or gender (ß=0.46, SE = 0.25, 95% CI = 0.961-2.612, χ2 = 3.25, p =.07, OR = 1.58) was found.

In summary, these findings indicate that the greater the number of symptoms COVID-19 patients experienced during the first 2 weeks of illness, the more likely they are to have long-term symptoms, which is in line with previous findings ^41^. This is also in line with more severe outcomes of hospitalized versus non-hospitalized COVID-19 patients ^29^.

